# Trends and Burden of Breast and Cervical Cancer in Bangladesh: A Data-Driven Analysis and Future Outlook

**DOI:** 10.64898/2025.12.05.25341679

**Authors:** Abdullah Al Rafi, Kashefatun Nur Orni, Tasnuba Ashraf Subah, Saima Akther, Ankita Acharjee, Imran Al Zuhayr, Debabrata Mallick

**Author notes:** **Correspondence:** Debabrata Mallick.

## Abstract

**Introduction:** Among women in Bangladesh, breast and cervical cancers remain the most frequently diagnosed malignancies. To strengthen early detection, In 2005, a national VIA-based cervical cancer screening initiative was launched, followed later by the addition of CBE for breast cancer and later integrated Clinical Breast Examination (CBE) for breast cancer screening. Since 2013, the Ministry of Health and Family Welfare (MOHFW) has implemented an electronic reporting system based on DHIS2 for aggregated health data, complemented by a case-based reporting stream established under the National Centre for Cervical and Breast Cancer Screening and Training (NCCBCST). This study reviews and evaluates VIA, colposcopy, and CBE screening data from 2024–2025 to assess the effectiveness, reliability, and overall performance of the electronic reporting system.

**Methods:** A descriptive approach was employed, using secondary data obtained from both the aggregated and case-based DHIS2 electronic databases. The study included women aged 30–60 years screened for cervical cancer using VIA and all women undergoing CBE across multiple levels of health facilities, including community clinics, upazila and district hospitals, medical colleges, tertiary hospitals, and BSMMU. Data from 2024–2025 were cleaned and analysed in Microsoft Excel to assess screening volumes, positivity rates, and division-wise variations. Aggregated and case-based datasets were compared to evaluate completeness, consistency, and functionality of the reporting system.

**Results:** Across 2024–2025, a total of 917,845 VIA tests and 964,228 CBE tests were performed nationwide. The national VIA-positivity rate was 9.7%, with the highest rates observed in Mymensingh (12.1%) and Rangpur (11.4%). A total of 20,819 colposcopy tests were conducted, showing a high national colposcopy-positivity rate of 147.2%, indicating a substantial number of referrals originating outside routine VIA screening. CBE positivity averaged 4.9%, with notable regional variation—highest in Rangpur (7.1%) and lowest in Barishal (2.9%). Screening participation was highest in the Dhaka division, whereas Sylhet consistently exhibited the lowest levels of service use. Aggregated and case-based reporting displayed strong alignment for key indicators, though referral pathway inconsistencies were observed.

**Conclusions:** Electronic reporting through DHIS2 both aggregated and case-based proved effective for monitoring cervical and breast cancer screening activities in Bangladesh, enabling large-scale data capture, geographic analysis, and programme oversight. However, substantial inter-divisional disparities, low screening uptake in specific regions, and referral-linkage gaps between VIA and colposcopy highlight areas for improvement. Improving quality control mechanisms, adopting unique identifiers, and reinforcing referral pathways could enhance overall equity and efficiency of the national cancer-prevention programme.

## Introduction

Currently, breast and cervical cancers represent the most common malignancies among women in Bangladesh and globally. According to recent cancer statistics, female breast cancer is the leading cancer among Bangladeshi women, while cervical cancer still contributes substantially to morbidity and mortality despite improvements in screening uptake **[1]**. In response to national commitments to strengthening women’s health services **[2]**, the Ministry of Health and Family Welfare (MOHFW) launched a nationwide cervical cancer screening programme in 2005 using Visual Inspection with Acetic Acid (VIA) for women aged 30–60 years **[3][4][5]**. For breast cancer, Clinical Breast Examination (CBE) was adopted as the primary early-detection strategy and integrated into the essential service package of routine health facilities, supported by technical guidance from the World Health Organization (WHO) **[6]**. To improve monitoring and data-driven programme management, the MOHFW introduced an electronic aggregated reporting system in 2013 using the District Health Information System version 2 (DHIS2), enabling monthly submission of population-level indicators from national to subnational health facilities **[7][8]**. DHIS2 has become widely used across low- and middle-income countries for public-health reporting because of its flexibility in data entry, validation, visualization, and routine analysis **[9][10]**. In Bangladesh, the establishment of the national centre for cervical and breast cancer screening Bangladesh at Bangabandhu Sheikh Mujib Medical University (BSMMU) further strengthened service quality through competency-based training, coordination, and expansion of VIA and CBE services to district and upazila levels **[11][12]**. Currently, two digital reporting streams aggregated DHIS2 data and case-based NCCBCST data—operate in parallel to support national surveillance. Programme dashboards show substantial screening volumes. For the 2025 case-based data collection system, a total of 408,828 women were enrolled, including 90,966 (22.3%) in community clinics and 317,862 (77.7%) in other health facilities. The system recorded 313,227 VIA screenings, with 11,319 women testing VIA-positive (3.6%). A total of 11,814 colposcopy tests were conducted, of which 4,922 (41.7%) were colposcopy-positive. Breast cancer screening data showed 333,484 CBE screenings, with 5,272 women (1.6%) testing CBE-positive. In the 2025 aggregated data collection system, 448,048 VIA screenings were reported, with 15,589 VIA-positive cases (3.5%). Additionally, 6,244 colposcopy tests were conducted, identifying 2,994 colposcopy-positive cases (48.0%). Breast cancer indicators included 459,256 CBE screenings, with 9,903 women (2.2%) testing CBE-positive**[13]**. For the 2024 case-based data collection system, a total of 1,069,260 women were enrolled, including 553,169 (51.7%) in community clinics and 516,091 (48.3%) in other health facilities. The system recorded 604,718 VIA screenings, with 17,138 women testing VIA-positive (2.8%). A total of

17,205 colposcopy tests were conducted, of which 6,860 (39.9%) were colposcopy-positive. Breast cancer screening data showed 630,745 CBE screenings, with 7,524 women (1.2%) testing CBE-positive. In the 2024 aggregated data collection system, 717,591 VIA screenings were reported, with 18,579 VIA-positive cases (2.6%). Additionally, 8,843 colposcopy tests were performed, identifying 4,244 colposcopy-positive cases (48.0%). Breast cancer indicators included 726,533 CBE screenings, with 10,958 women (1.5%) testing CBE-positive. **[13]**. These routine data allow assessment of screening coverage, positivity rates, referral completion for colposcopy, and overall data-system performance. Despite long-term electronic reporting, systematic evaluation of these DHIS2-based datasets particularly completeness, accuracy, and programme utility remains limited. Therefore, the objective of this paper is to summarise and analyse VIA and CBE screening data from 2024-2025, using both aggregated and case-based DHIS2 systems, to determine the effectiveness and usefulness of electronic data collection for monitoring cervical and breast cancer screening outcomes in Bangladesh.

## Methods

This study employed a descriptive design using secondary data extracted from the electronic repository of Bangladesh’s national cervical and breast cancer screening programme. Both aggregated and case-based data were obtained from the District Health Information System version 2, which serves as the national platform for routine reporting. Monthly submissions from health facilities across all administrative levels contributed to the datasets analysed for the years 2024 and 2025. The study population consisted of women aged 30–60 years who underwent cervical cancer screening using Visual

Inspection with Acetic Acid (VIA) and breast cancer screening using Clinical Breast Examination (CBE). Screening services were delivered through a diverse range of health facilities, including Specialized Hospitals, Medical College Hospitals, District/General Hospitals, Maternal & Child Welfare Centres (MCWCs), Sadar and Upazila Health Complexes (UHCs), 10/20/31/50-Bed Hospitals, Postgraduate Institutes & Hospitals, District-Level Hospitals, Community Clinics, and medical universities such as Bangabandhu Sheikh Mujib Medical University (BSMMU). Both national- and subnational-level facilities contributed to the datasets. Data were extracted from DHIS2 and cleaned using Microsoft Excel to remove duplications, inconsistencies, and missing entries. Variables included geographic identifiers (division and district), reporting month and year, facility type, total enrollment, VIA screening numbers and positivity rates, colposcopy referral and positivity, and CBE screening and positivity. For 2025, the case-based dataset included 408,828 total enrollments, of which 90,966 (22.3%) were from community clinics and 317,862 (77.7%) from other health facilities. It also contained 313,227 VIA screenings (3.6% VIA-positive), 11,814 colposcopy tests (41.7% positive), and 333,484 CBE screenings (1.6% positive). The aggregated dataset for 2025 included 448,048 VIA screenings (3.5% VIA-positive), 6,244 colposcopy tests (48.0% positive), and 459,256 CBE screenings (2.2% positive). For 2024, the case-based dataset included 1,069,260 enrolled women, with 604,718 VIA screenings (2.8% VIA-positive), 17,205 colposcopy tests (39.9% positive), and 630,745 CBE screenings (1.2% positive). The 2024 aggregated dataset documented 717,591 VIA screenings (2.6% VIA-positive), 8,843 colposcopy tests (48.0% positive), and 726,533 CBE screenings (1.5% positive). Both case-ba sed and aggregated data we re available for all eight divisions—Chattogram, Barishal, Dhaka, Khulna, Mymensingh, Rajshahi, Rangpur, and Sylhet. Descriptive analysis was conducted using Microsoft Excel to examine the distribution of VIA and CBE screenings, positivity rates, and colposcopy referral outcomes across facility types and geographic regions. Comparisons between case-based and aggregated reporting streams were performed to assess consistency, completeness, and coverage of cervical and breast cancer screening services. The analysis provided insight into the performance of electronic data reporting systems and their role in monitoring national screening outcomes.

## Results

Figure **1.1** Distribution of VIA Tests and VIA-Positive Cases (2024–2025) Figure 1.1 illustrates the division-wise cumulative number of VIA tests performed between 2024 and 2025 in Bangladesh. A total of 917,845 VIA tests were conducted across the eight administrative divisions. The highest number of VIA tests was performed in the Dhaka division (247,037 tests; 26.9%), followed by the Rajshahi (143,659 tests; 15.6%) and Chattogram (136,809 tests; 14.9%) divisions. The lowest number of tests was done in Sylhet (59,527 tests; 6.5%). The corresponding distribution of VIA-positive cases shows a total of 28,457 women who tested positive during the same period. Dhaka division again contributed the highest proportion (7,751 cases; 27.2%), followed by Rajshahi (4,959 cases; 17.4%) and Khulna (4,624 cases; 16.3%). The most minor proportion of VIA-positive cases was observed in Sylhet (2,246 cases; 7.9%). Overall, the national VIA-positivity rate was 9.7%.

**Fig. 1.1.**
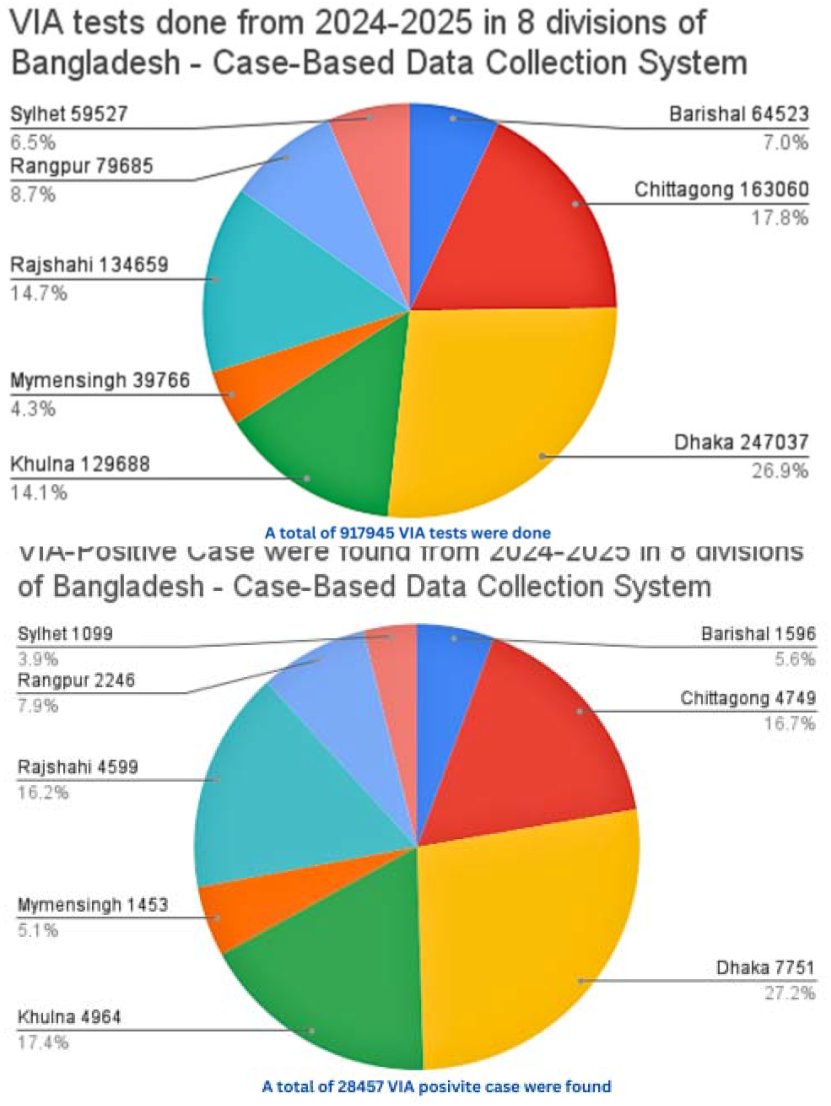
Division-wise distribution of number of VIA tests done and VIA positive case found from 2024 to 2025 in 8 divisions of Bangladesh

Figure **1.2** Distribution of Colposcopy Tests and Colposcopy-Positive Cases (2024–2025) Figure 1.2 shows the distribution of colposcopy tests performed across the eight divisions. A total of 20,819 colposcopy tests were conducted during 2024– 2025. The highest number was performed in Dhaka division (9,362 tests; 45.0%), followed by Rajshahi (3,475 tests; 16.7%) and Chattogram (3,469 tests; 16.7%). The lowest number was in Sylhet (1,059 tests; 5.1%). The distribution of colposcopy-positive cases reveals 13,782 positive cases.

**Fig. 1.2.**
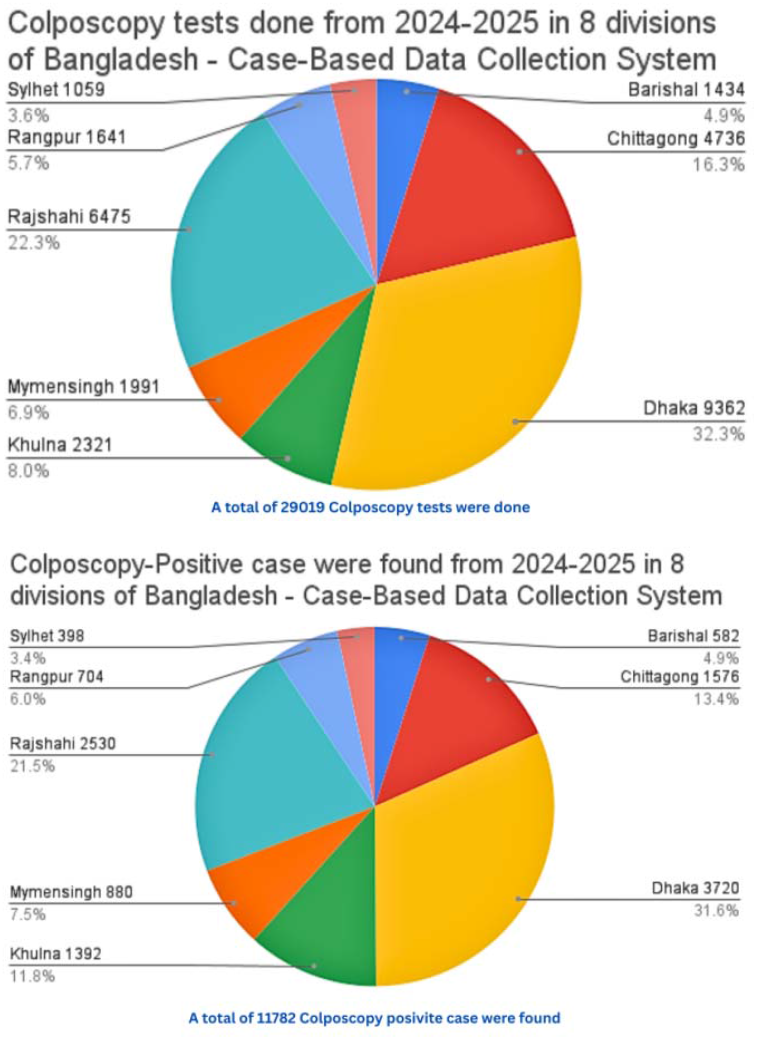
Division-wise distribution of number of Colposcopy tests done and VIA positive case found from 2024 to 2025 in 8 divisions of Bangladesh

Women nationwide. Dhaka division had the highest share (3,720 cases; 27.0%), followed by Rajshahi (3,312 cases; %) and Khulna (1,392 cases; 10.1%). The lowest proportions were found in Sylhet (398 cases; 2.9%) and Barishal (582 cases; 4.2%). The national colposcopy-positivity rate was 147.2%, suggesting many VIA-positive referrals arrived from outside the routine screening system.

Figure **1.3** Distribution of CBE Tests and CBE-Positive Cases (2024–2025) Figure 1.3 summarizes the distribution of CBE (Clinical Breast Examination) tests conducted across the divisions between 2024 and 2025. A total of 964,228 CBE tests were recorded. The most significant number was done in Dhaka division (257,093 tests; 26.7%), followed by Rajshahi (142,645 tests; 14.8%), and Chattogram (172,632 tests; 17.9%). The lowest number of tests was performed in Sylhet (51,488 tests; 5.3%). The total number of CBE-positive cases identified was 12,976. Dhaka division reported the highest proportion (3,511 cases; 27.1%), followed by Rajshahi (2,243 cases; 17.3%) and Chattogram (1,696 cases; 13.1%). The lowest number of positive instances occurred in Barishal (2,094 cases; 16.1%) and Sylhet (1,314 cases; 10.1%). The national CBE-positivity rate was 4.9%.

**Fig. 1.3.**
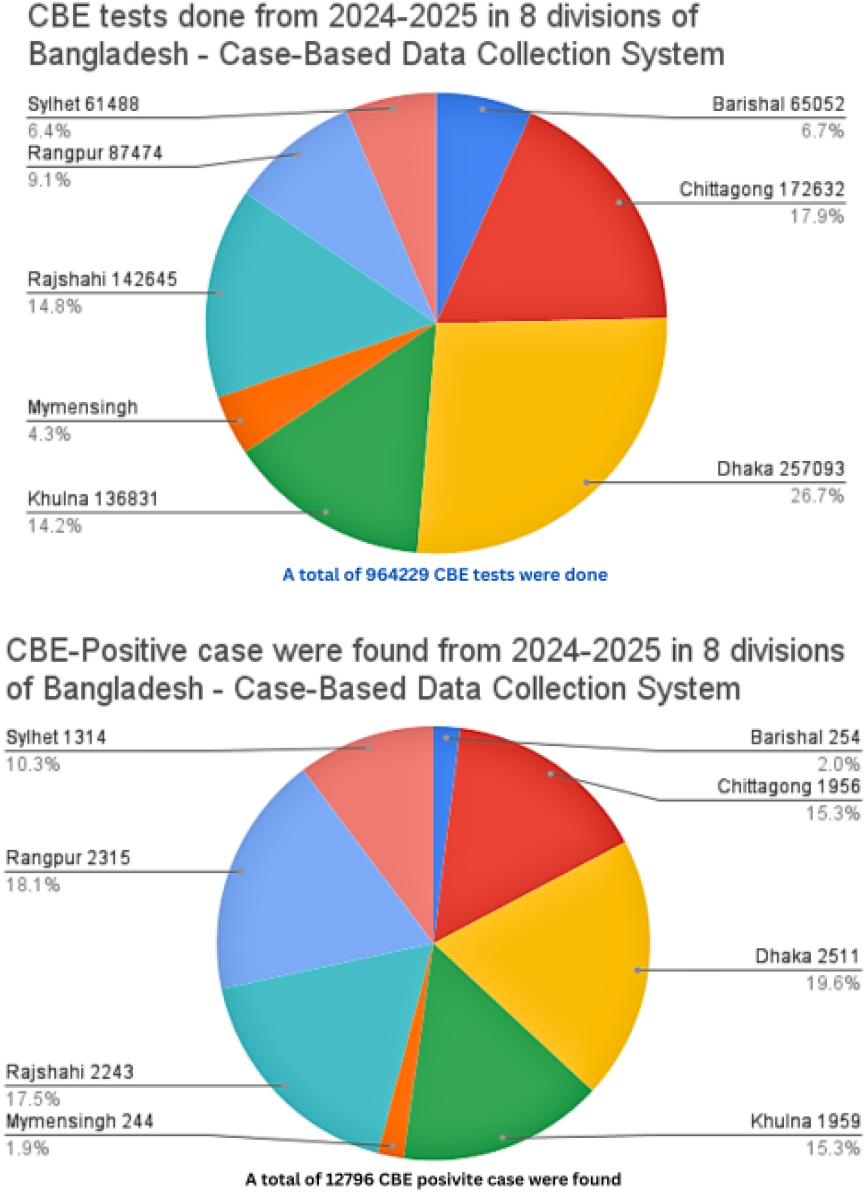
Division-wise distribution of number of Colposcopy tests done and Colposcopy positive case found from 2024 to 2025 in 8 divisions of Bangladesh

Figure **2.1** Division-wise VIA-Positivity Rate (2024–2025) Figure 2.1 illustrates the Variation in VIA-positivity rates across the eight administrative divisions of Bangladesh from 2024 to 2025. The national VIA-positivity rate during this period was 9.7%. The highest VIA-positivity was observed in Mymensingh division (12.1%), followed closely by Rangpur (11.4%), Khulna (11.2%), and Rajshahi (10.9%). Moderate positivity rates were recorded in Dhaka (9.7%) and Chattogram (9.5%). The lowest rates were seen in Barishal (8.0%) and Sylhet (5.9%). Overall, the curve shows substantial inter-divisional variability, with Mymensingh and Rangpur contributing the highest positivity burden.

**Fig. 2.1.**
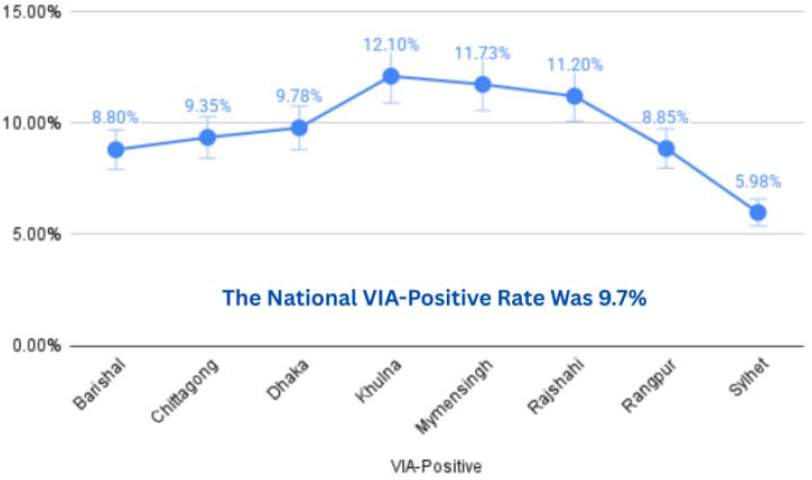
Division-wise VIA-positivity rate from 2024 to 2025 in 8 divisions of Bangladesh

Figure **2.2** Division-wise Colposcopy-Positivity Rate (2024– 2025) Figure 2.2 demonstrates the colposcopy-positivity rate among women referred for the procedure across eight divisions. The national colposcopy-positivity rate during the study period was exceptionally high at 147.2%, indicating that many women underwent colposcopy without prior VIA screening or were referred from multiple sources. The highest positivity rate was reported in Mymensingh division (203.8%), followed by Barishal (163.6%), Sylhet (157.9%), and Dhaka (158.1%). Lower positivity proportions were observed in Rajshahi (129.4%), Chattogram (115.2%), and Khulna (141.4%). The sharp peak in Mymensingh suggests a heavier influx of colposcopy referrals relative to the number of VIA-positive.

**Fig. 2.2.**
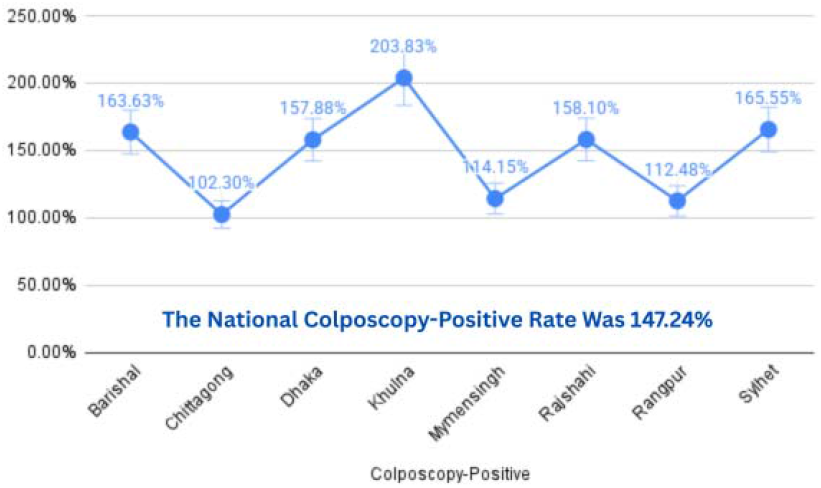
Division-wise Colposcopy-positivity rate from 2024 to 2025 in 8 divisions of Bangladesh

### Case-Based Data Collection System (2024-2025)

Figure **2.3** Division-wise CBE-Positivity Rate (2024–2025) Figure 2.3 presents the distribution of CBE-positivity rates across the eight divisions of Bangladesh during 2024–2025. The national average CBE-positivity rate was 4.9%. The highest CBE-positivity rate was recorded in Rangpur division (7.1%), followed by Rajshahi (6.0%), Khulna (5.6%), and Chattogram (4.5%). Moderate rates were found.

In Mymensingh (3.5%) and Dhaka (3.5%), the lowest rates were reported in Barishal (2.9%) and Sylhet (3.8%). The data indicate that breast-related abnormality detection varies across divisions, with Rangpur showing a considerably higher positivity burden.

**Fig. 2.3.**
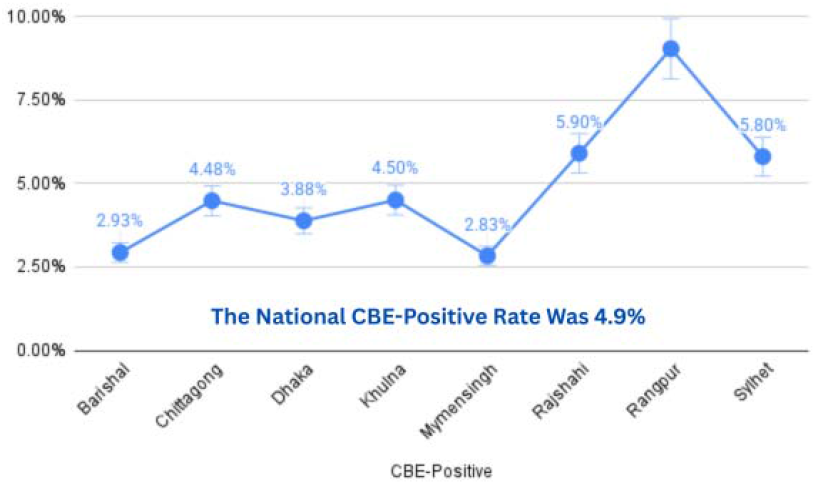
Division-wise CBE-positivity rate from 2024 to 2025 in 8 divisions of Bangladesh

## Discussions

This analysis of DHIS2-based aggregated and case-based data from 2024–2025 provides an updated overview of the performance of Bangladesh’s national cervical and breast cancer screening programmes. The findings demonstrate substantial geographic Variation in service uptake and positivity rates, highlighting both the strengths and persisting gaps in early-detection services across the country. These observations align with national strategic priorities for cancer prevention and the evolving burden of cervical and breast cancer in Bangladesh **[1][2][3][4]**.

### Sustained Screening Activity but Uneven Uptake

Sustained Screening Activity but Uneven Uptake. A total of 917,845 VIA tests and 964,228 CBE tests were performed over the two-year period, indicating high engagement of health facilities and a functional electronic reporting system, consistent with previous DHIS2 implementation reports from Bangladesh **[10][11]**. Similar to earlier national assessments, Dhaka division contributed the largest share of total screenings for both VIA and CBE, reflecting its population density and concentration of tertiary health facilities **[13]**. Conversely, Sylhet consistently recorded the lowest screening volumes, reflecting long-documented challenges in health-seeking behaviour and service utilization in this division, which mirrors findings from previous national reports **[11][13]**. These patterns reaffirm regional health inequities previously reported by DGHS and national surveys **[3][4]**.

### Geographic Variation in VIA Positivity

The national VIA positivity rate (9.7%) was notably higher than the long-term average of 3.3–3.8% reported between 2014 and 2022 **[11]**. The divisional map reveals that Mymensingh, Rangpur, Khulna, and Rajshahi showed higher VIA positivity (10–12%), whereas Sylhet and Barishal showed the lowest positivity (5–8%). District-level patterns reinforce these disparities. These differences may reflect: True Variation in disease burden, influenced by early marriage, multiparity, and limited HPV vaccination coverage, as documented in global and national cervical cancer epidemiology reports **[1][2]**; Differences in provider performance, especially related to VIA interpretation quality, an issue highlighted in WHO’s cervical cancer screening guidelines **[5]**; Referral concentration, where districts with strong referral hospitals receive VIA-positive women from neighbouring areas. Such findings underscore the need for periodic quality assurance, standardized VIA training, and targeted community mobilization in low-uptake regions, as emphasized in Bangladesh’s national cervical cancer strategy **[4]**.

### Exceptionally High Colposcopy Positivity

A striking observation in this dataset is the very high colposcopy positivity rate (147.2%), with divisional rates exceeding 150% in several regions. These values confirm that: Many women underwent colposcopy without prior VIA testing, likely due to symptomatic presentation or direct referrals from outpatient departments, a trend previously noted by NCCBCST and BSMMU reports**[12][13]**. Regional referral centres receive patients from multiple districts, inflating positivity ratios. Backlogs from previous years, including the post-COVID recovery period, may have increased referrals for clinically suspicious cases. While the high positivity may reflect effective detection of high-risk cases, it also highlights gaps in the referral chain and incomplete linkage between VIA and colposcopy services. Strengthening the continuum of care from VIA to colposcopy to treatment is essential and has been strongly recommended in WHO guidelines **[5]** and national programme documents **[4]**.

### Breast Cancer Screening: Lower Positivity but Marked Regional Differences

CBE positivity (4.9%) was lower than VIA and colposcopy positivity, consistent with global observations that CBE identifies fewer abnormalities than cervical precancer screening **[6][7]**. However, the district-level maps revealed notable regional Variation. Rangpur (7.1%) and Rajshahi (6.0%) had the highest positivity, while Barishal and Sylhet recorded the lowest. These differences may reflect: Greater breast symptom awareness in high-positivity divisions, Variation in examiner skill and CBE technique, Differences in the age distribution of screened women. Given the rising burden of breast cancer in Bangladesh **[2][6]**, targeted provider training and standardized protocols for early detection are needed.

### Implications for Data Quality and the Electronic Reporting System

The strong alignment between aggregated and case-based reporting for most indicators suggests increasing reliability of the DHIS2 system nationally. The steady increase in facility participation demonstrates growing institutional acceptance of digital reporting platforms, consistent with global DHIS2 documentation and implementation experiences **[8][9]**. However, inconsistencies such as colposcopy positivity exceeding 100% highlight limitations in the referral tracking mechanism. These findings underscore the need for: Improved linkage between VIA and colposcopy modules, Unique patient identifiers for longitudinal follow-up, and Integration of DHIS2 with NCCBCST case-based data, as recommended in recent national dashboards **[12][13]**. Despite these gaps, Bangladesh’s combined aggregated and case-based reporting platforms remain among the most comprehensive cancer screening information systems in South Asia **[11]**.

### Comparison With International Data

The national VIA positivity rate (9.7%) and CBE positivity rate (4.9%) fall within the range reported in other LMICs, although overall screening coverage remains lower than in countries such as Thailand (∼54%) and England (∼78%), as reported by global cancer observatories and WHO screening comparisons **[1][5]**. Nevertheless, coverage exceeds that of several Sub-Saharan African countries (3–7%) **[5]**, reflecting both progress and the existing need for rapid scale-up to achieve the WHO’s cervical cancer elimination targets.

### Strengths and Public Health Importance

This study is one of the first to combine divisional and district-level maps for VIA, colposcopy, and CBE positivity using DHIS2 data. The geographic visualization clearly identifies areas requiring: Increased outreach (e.g., Sylhet, Barishal), Enhanced VIA provider training (e.g., Mymensingh, Rangpur), Strengthened referral management (e.g., Barishal, Sylhet high colposcopy positivity).

These insights are directly applicable to national programme implementation and align with the direction of recent MOHFW policy documents **[3][4][13]**.

**Figure 3.1** shows the distribution of Visual Inspection with Acetic Acid (VIA) positivity rates across divisions. Highest Rates (High ≥10% : Khulna (12.1%) and Mymensingh (11.73%) reported the highest positivity rates.Moderate Rates (Moderate 5–10%): Rajshahi (11.2% - note: this is labeled moderate but is >10%, potentially an error in the legend/coloring used in the image), Dhaka (9.78%), Chittagong (9.35%), Rangpur (8.85%), Barishal (8.0%), and Sylhet (5.98%) fall into the moderate category.The data suggests significant variation in the prevalence of VIA-detectable abnormalities across the country.

**Fig. 3.1.**
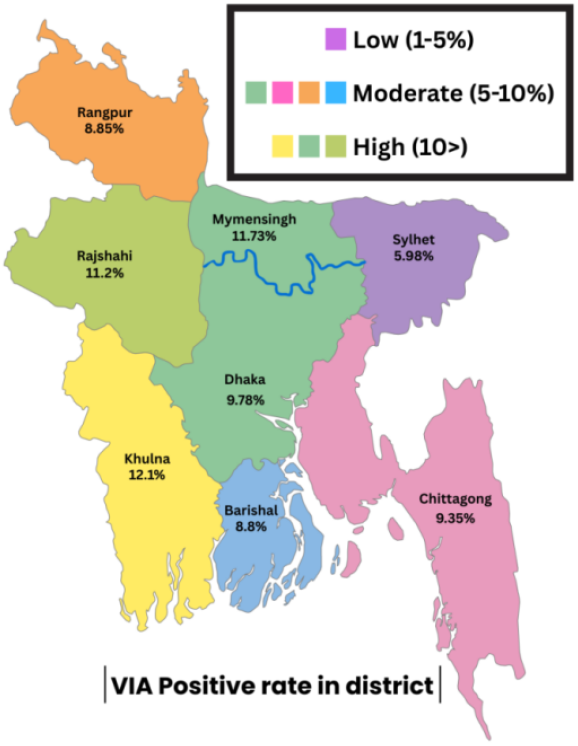
Division-wise distribution of cumulative VIA-positive rates in map of Bangladesh over 2-years (2024 to 2025)

**Figure 3.2** presents the distribution of Colposcopy positivity rates, which are significantly higher than the other two tests, suggesting a high rate of abnormalities confirmed by colposcopy, or possibly a high rate of false positives from the preceding screening test (not provided here). The legend is scaled differently: Low (1-100%), Moderate (100-150%), High (≥150%).Highest Rates (≥150%): Khulna (203.63%), Chittagong (202.3%), Rajshahi (158.2%), and Dhaka (157.88%) reported the highest rates.Moderate Rates (Moderate 100–150%): Barishal (143.63%), Sylhet (145.55%), Mymensingh (114.15%), and Rangpur (112.48%) fall into the moderate range.All divisions show very high Colposcopy-positivity rates, which are all over 100%.

**Fig. 3.2.**
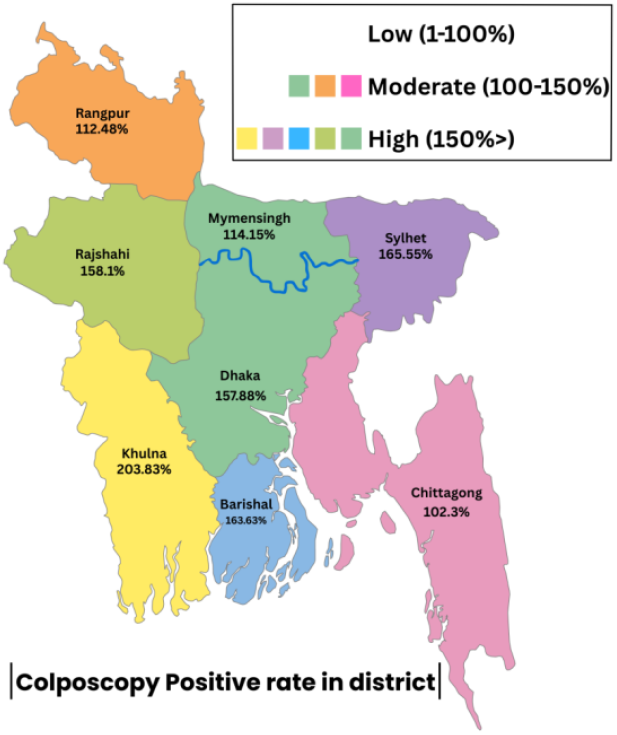
Division-wise distribution of cumulative,colposcopy positive rates in map of Bangladesh over 2-years (2024 to 2025)

**Figure 3.3** presents the distribution of Clinical Breast Examination (CBE) positivity rates. The legend categorizes rates as Low (1-5), Moderate (5-10%), and High (≥10%).Highest Rates (High ≥10%): Rangpur (9.03% - note: this is labeled High in the map but falls in the Moderate category based on the legend’s range) reported the highest rate.Moderate Rates (Moderate 5–10%): Rajshahi (5.9%) and Sylhet (5.8%) show moderate rates of CBE-detectable breast abnormalities.Lowest Rates (Low 1–5%): Dhaka (3.88%), Mymensingh (2.63%), Khulna (4.5%), Chittagong (4.48%), and Barishal (2.93%) show the lowest positivity rates.The data indicates a relatively low overall rate of CBE-positivity compared to the other two tests, with Rangpur showing the highest burden.

**Fig. 3.3.**
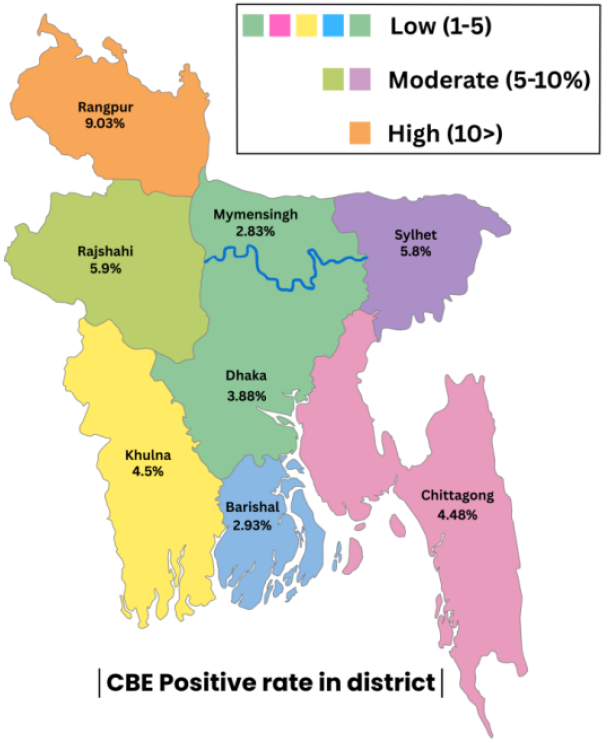
Division-wise distribution of CBE-positivity rates in map of Bangladesh over 2-years (2024 to 2025)

## Conclusions

Overall, Bangladesh’s electronic cervical and breast cancer screening system demonstrates substantial progress in coverage, data availability, and geographic monitoring. However, regional disparities, low screening uptake in certain divisions, and inconsistencies in the VIA– colposcopy referral pathway require urgent attention. Strengthening quality assurance, improving patient tracking, and expanding targeted outreach will be critical to achieving equitable cancer prevention nationwide.

## Data Availability

All data produced in the present work are contained in the manuscript.

https://cancer-dashboard.mohfw.gov.bd/cervical-breast/

## Notes

### Competing Interest Statement

The authors have declared no competing interest.

### Funding Statement

This study did not receive any funding

### Author Declarations

This study used ONLY openly available human data that were originally located at : https://cancer-dashboard.mohfw.gov.bd/cervical-breast/

